# Epidemiology of Atrial Fibrillation in Colombia 2010-2015

**DOI:** 10.1101/2023.06.27.23291899

**Authors:** Pablo Andrés Miranda-Machado, Roberto Carlos Guzman-Saenz

**Affiliations:** Epidemiologia, ALZAK Foundation. Cartagena, Colombia; Universidad Norte. Barranquilla, Colombia

**Keywords:** atrial fibrillation, epidemiology, prevalence, incidence, demand for care, Colombia

## Abstract

Atrial Fibrillation (AF) is a common cardiac arrhythmia. Its prevalence worldwide has doubled in the last decade due to increased capacity for diagnosis and management of heart disease and chronic non-cardiac. The purpose of this study was to estimate the prevalence, incidence and demand for treatment of AF in Colombia during the period 2010 to 2015.

## Introduction

Atrial Fibrillation (AF) is the most frequent and most important cardiac arrhythmia of the clinical practice in the adult population. Both due to its incidence, which increases with age, and due to its morbidity and mortality given its ability to cause cardioembolic and cerebrovascular events (1). The main risk factor for developing AF is the age of the patient. The older the age the higher the incidence. Arterial hypertension, diabetes, heart failure are also found as the main risk factors. Other related risk factors are hyperthyroidism, maternal obesity and high birth weight (greater than 3.5 kg), fatty liver, consumption of alcohol and hallucinogenic drugs. The important thing is that these risk factors are potentially modifiable, by changing lifestyles and therapeutic interventions (2-8). It requires the efforts of the primary care physician for prompt recognition through basic tests as the electrocardiogram (EKG)(9) in older adults with risk factors (high blood pressure (hypertension), diabetes mellitus (DM), cerebrovascular disease, heart failure, valvular disease, coronary heart disease, etc). and rigorous screening in second and third level centers in patients with acute cerebrovascular events with medium complexity tests such as 24-hour ambulatory EKG monitoring, continuous in-hospital EKG monitoring and echocardiography (10,11). This will allow a timely start treatment to control heart rate and / or control the pace, and above all prevent stroke with oral anticoagulation (12,13).

The global prevalence of AF is 1-2%. Its increases with age and it is estimated that the risk of developing this arrhythmia after 40 years of age is 25%. It occurs more in men than in women and is identified in 1 in 20 patients who present cerebrovascular events (14). The global prevalence of 33.5 million patients was estimated for 2010, in the United States, with a prevalence of 5.0% of the population. Approximately 5.2 million patients, with an annual incidence of 1.2 million patients. For Latin America the prevalence figure was estimated at 1.5 to 2.0% of the population, approximately 7 million patients, in Brazil alone it is estimated at 1.5 million patients.

The prevalence of cerebrovascular disease (CVA) in Latin America ranges between 5.7 and 7.7 / 1000 inhabitants, of which about 10% of all hospitalized patients in Mexico and Brazil documented atrial fibrillation, and 8% in Argentina. In China, in 2008, a prevalence of 0.9% of the population over 30 years of age was estimated, from which a risk of cerebrovascular events of 12.5% was calculated with respect to 2.5% of the general population without fibrillation atrial (AF) (1,2,15–17). The purpose of this study was to estimate the prevalence, incidence and demand for AF care in Colombia between 2010 and 2015.

## Methods

A cross-sectional study was carried out. The total number of people that attended nationwide who had a diagnosis of AF (ICD-10 I48X) was included in the Database of the Social Protection Information System (SISPRO) for the period between 2010 and 2015. To estimate the prevalence, all types of diagnosis were included and new confirmed diagnoses were included to estimate the incidence. For the adjustments of the prevalence and incidence, the estimates of the national population of the National Department of Statistics (DANE) for the period between 2010 and 2015 were included. To estimate the prevalence of demand for care, the total number of people served at the nationwide registration in the SISPRO Database, between the period 2010 and 2015 was included. The qualitative and quantitative variables were expressed in percentages, and means with standard deviations respectively. All estimates were adjusted for age and sex.

## Results

Between 2010 and 2015, a total of 112,353 people attended with a diagnosis of AF, were identified for an average of 22,470 + 6,413 patients with a diagnosis of AF per year. The prevalence of AF estimated in 2010 and 2015 was 22.6 and 51.4 per 100,000, respectively; and more than 60% of the patients were older than 65 years. The prevalence of AF in women and men in 2010 was 22.6 and 22.5 per 100,000, respectively. In 2015, the prevalence of AF in women and men was 51.2 and 51.5 per 100,000. In patients between 40-45 years, the prevalence of AF was higher in men in both 2010 and 2015 (2010 men 10.1 per 100,000 vs. women 5.8 per 100,000, 2015 men 16.2 per 100,000 vs. women 9.9 per 100,000) (Table 1-5). A total of 173,784 people treated with a newly confirmed diagnosis of AF were identified between 2010 and 2015, for an average of 2,897 + 887 patients with a new diagnosis of AF each year. The estimated incidence of AF in 2010 and 2015 was 4.1 and 8.0 per 100,000 respectively; and more than 66% of the patients were older than 65 years. The incidence of AF in women and men in 2010 was 4.2 and 3.9 per 100 thousand, respectively. In 2015, the incidence of AF in women and men in 2015 was 8.1 and 7.8 per 100,000. In patients between 40-45 years, the incidence of AF was similar between men and women in both 2010 and 2015 (2010 men 1.2 per 100,000 vs. women 1.3 per 100,000, 2015 men 1.9 per 100,000 vs. women 1.0 per 100 thousand) (Table 1,2, 6-8).

**Table 1.**
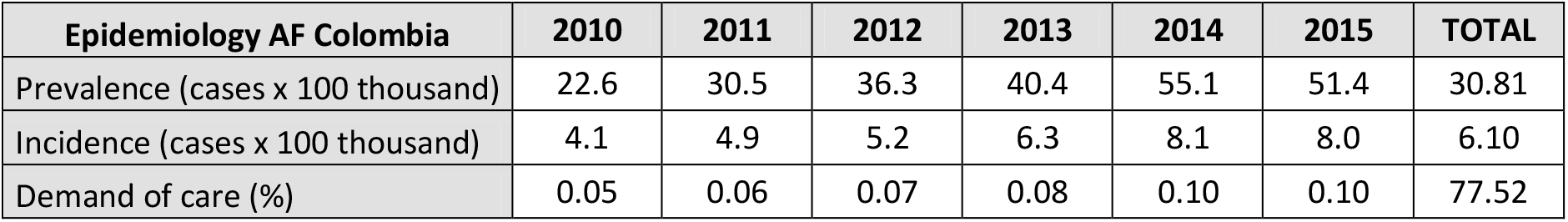
Epidemiology of AF in Colombia 2010-2015. Source: SISPRO-DANE.

**Table 2.** Total population, attended population and cases of AF in Colombia 2010-2015. Source: SISPRO-DANE.

| **Total population, attended and cases** | **2010** | **2011** | **2012** | **2013** | **2014** | **2015** |
| --- | --- | --- | --- | --- | --- | --- |
| Total population | 45509584 | 46044601 | 46581823 | 47121089 | 47661787 | 48203405 |
| Attended population | 18637420 | 20797416 | 22421024 | 22273815 | 26289936 | 24354571 |
| Prevalent cases AF | 10303 | 14038 | 16901 | 19073 | 27231 | 24807 |
| Incident cases AF | 1870 | 2254 | 2423 | 2973 | 3989 | 3876 |

**Table 3.** Prevalent cases of AF in Colombia 2010-2015 for ages. Source: SISPRO.

| **Ages** | **2010** | **2011** | **2012** | **2013** | **2014** | **2015** |
| --- | --- | --- | --- | --- | --- | --- |
| De 0 a 4 years | 15 | 20 | 31 | 19 | 85 | 45 |
| De 05 a 09 years | 9 | 10 | 12 | 12 | 98 | 24 |
| De 10 a 14 years | 14 | 21 | 8 | 17 | 73 | 35 |
| De 15 a 19 years | 55 | 30 | 48 | 52 | 151 | 68 |
| De 20 a 24 years | 61 | 71 | 79 | 73 | 199 | 148 |
| De 25 a 29 years | 91 | 108 | 132 | 123 | 219 | 128 |
| De 30 a 34 years | 117 | 148 | 163 | 187 | 333 | 232 |
| De 35 a 39 years | 146 | 170 | 202 | 189 | 371 | 285 |
| De 40 a 44 years | 230 | 280 | 332 | 340 | 481 | 376 |
| De 45 a 49 years | 384 | 453 | 568 | 570 | 778 | 685 |
| De 50 a 54 years | 583 | 735 | 819 | 893 | 1,142 | 1,036 |
| De 55 a 59 years | 733 | 1,026 | 1,161 | 1,335 | 1,808 | 1,645 |
| De 60 a 64 years | 1,022 | 1,363 | 1,604 | 1,823 | 2,392 | 2,285 |
| De 65 a 69 years | 1,269 | 1,716 | 1,996 | 2,293 | 3,234 | 3,143 |
| De 70 a 74 years | 1,549 | 2,175 | 2,596 | 2,819 | 3,983 | 3,671 |
| De 75 a 79 years | 1,736 | 2,298 | 2,880 | 3,282 | 4,562 | 4,172 |
| De 80 years or older | 2,551 | 3,702 | 4,726 | 5,438 | 8,023 | 7,489 |
| **Total** | **10,303** | **14,038** | **16,902** | **19,073** | **27,231** | **24,826** |

**Table 4.** Prevalent cases of AF in Colombia (Female) 2010-2015 for ages. Source: SISPRO.

| **Ages** | **2010** | **2011** | **2012** | **2013** | **2014** | **2015** |
| --- | --- | --- | --- | --- | --- | --- |
| De 0 a 4 years | 8 | 9 | 16 | 7 | 44 | 16 |
| De 05 a 09 years | 3 | 6 | 7 | 7 | 50 | 11 |
| De 10 a 14 years | 8 | 10 | 3 | 8 | 34 | 15 |
| De 15 a 19 years | 25 | 18 | 29 | 29 | 81 | 35 |
| De 20 a 24 years | 25 | 26 | 40 | 39 | 113 | 77 |
| De 25 a 29 years | 21 | 34 | 44 | 36 | 108 | 52 |
| De 30 a 34 years | 37 | 35 | 69 | 64 | 147 | 82 |
| De 35 a 39 years | 45 | 68 | 80 | 72 | 172 | 119 |
| De 40 a 44 years | 88 | 103 | 132 | 128 | 211 | 150 |
| De 45 a 49 years | 166 | 193 | 237 | 233 | 345 | 290 |
| De 50 a 54 years | 254 | 300 | 391 | 399 | 514 | 445 |
| De 55 a 59 years | 350 | 476 | 528 | 624 | 854 | 750 |
| De 60 a 64 years | 442 | 598 | 708 | 808 | 1,101 | 1,014 |
| De 65 a 69 years | 546 | 772 | 917 | 1,037 | 1,479 | 1,385 |
| De 70 a 74 years | 775 | 1,063 | 1,231 | 1,392 | 1,941 | 1,731 |
| De 75 a 79 years | 989 | 1,254 | 1,614 | 1,796 | 2,453 | 2,249 |
| De 80 years or older | 1,570 | 2,221 | 2,768 | 3,243 | 4,705 | 4,389 |
| **Total** | **5,223** | **7,035** | **8,585** | **9,725** | **14,003** | **12,514** |

**Table 5.** Prevalent cases of AF (Male) in Colombia 2010-2015 for ages. Source: SISPRO.

| **Ages** | **2010** | **2011** | **2012** | **2013** | **2014** | **2015** |
| --- | --- | --- | --- | --- | --- | --- |
| De 0 a 4 years | 6 | 11 | 14 | 12 | 41 | 29 |
| De 05 a 09 years | 4 | 4 | 3 | 5 | 46 | 11 |
| De 10 a 14 years | 6 | 11 | 5 | 7 | 34 | 13 |
| De 15 a 19 years | 29 | 11 | 18 | 22 | 70 | 33 |
| De 20 a 24 years | 36 | 45 | 39 | 34 | 86 | 71 |
| De 25 a 29 years | 70 | 74 | 88 | 87 | 111 | 76 |
| De 30 a 34 years | 80 | 113 | 94 | 123 | 186 | 150 |
| De 35 a 39 years | 101 | 102 | 122 | 117 | 199 | 166 |
| De 40 a 44 years | 142 | 177 | 200 | 212 | 270 | 226 |
| De 45 a 49 years | 217 | 258 | 331 | 337 | 433 | 395 |
| De 50 a 54 years | 329 | 434 | 427 | 493 | 627 | 591 |
| De 55 a 59 years | 383 | 550 | 633 | 711 | 954 | 895 |
| De 60 a 64 years | 579 | 764 | 896 | 1,014 | 1,290 | 1,270 |
| De 65 a 69 years | 722 | 941 | 1,077 | 1,255 | 1,753 | 1,757 |
| De 70 a 74 years | 769 | 1,110 | 1,362 | 1,425 | 2,039 | 1,935 |
| De 75 a 79 years | 745 | 1,040 | 1,263 | 1,482 | 2,105 | 1,921 |
| De 80 years or older | 979 | 1,478 | 1,952 | 2,189 | 3,311 | 3,095 |
| **Total** | **5,065** | **6,986** | **8,298** | **9,333** | **13,204** | **12,289** |

**Table 6.** Incident cases of AF in Colombia 2010-2015 for ages. Source: SISPRO.

| **Ages** | **2010** | **2011** | **2012** | **2013** | **2014** | **2015** |
| --- | --- | --- | --- | --- | --- | --- |
| De 0 a 4 years | 4 | 3 | 4 | 1 | 1 | 1 |
| De 05 a 09 years | 1 |  | 1 |  | 2 | 5 |
| De 10 a 14 years | 3 | 3 |  | 3 |  | 5 |
| De 15 a 19 years | 13 | 4 | 10 | 9 | 15 | 9 |
| De 20 a 24 years | 6 | 5 | 9 | 11 | 19 | 13 |
| De 25 a 29 years | 15 | 12 | 24 | 11 | 15 | 18 |
| De 30 a 34 years | 18 | 27 | 25 | 19 | 30 | 36 |
| De 35 a 39 years | 32 | 22 | 34 | 27 | 55 | 41 |
| De 40 a 44 years | 38 | 57 | 50 | 52 | 57 | 42 |
| De 45 a 49 years | 77 | 88 | 95 | 90 | 105 | 101 |
| De 50 a 54 years | 104 | 105 | 117 | 147 | 166 | 159 |
| De 55 a 59 years | 136 | 176 | 178 | 194 | 255 | 231 |
| De 60 a 64 years | 172 | 220 | 246 | 284 | 354 | 329 |
| De 65 a 69 years | 241 | 256 | 276 | 382 | 497 | 505 |
| De 70 a 74 years | 276 | 352 | 351 | 439 | 639 | 582 |
| De 75 a 79 years | 318 | 398 | 415 | 531 | 636 | 648 |
| De 80 years or older | 448 | 550 | 614 | 797 | 1,199 | 1,228 |
| **Total** | **1,870** | **2,254** | **2,423** | **2,973** | **3,989** | **3,876** |

**Table 7.** Incident cases of AF (Female) in Colombia 2010-2015 for ages. Source: SISPRO.

| **Ages** | **2010** | **2011** | **2012** | **2013** | **2014** | **2015** |
| --- | --- | --- | --- | --- | --- | --- |
| De 0 a 4 years | 2 | 1 |  |  | 1 | 1 |
| De 05 a 09 years |  |  | 1 |  | 2 | 2 |
| De 10 a 14 years | 1 | 2 |  | 1 |  | 1 |
| De 15 a 19 years | 4 | 1 | 8 | 5 | 9 | 5 |
| De 20 a 24 years | 4 | 3 | 6 | 5 | 10 | 6 |
| De 25 a 29 years | 5 | 3 | 11 | 3 | 5 | 6 |
| De 30 a 34 years | 9 | 6 | 14 | 5 | 7 | 6 |
| De 35 a 39 years | 10 | 7 | 15 | 8 | 18 | 19 |
| De 40 a 44 years | 20 | 25 | 27 | 19 | 20 | 15 |
| De 45 a 49 years | 38 | 42 | 46 | 35 | 52 | 36 |
| De 50 a 54 years | 47 | 44 | 61 | 58 | 79 | 63 |
| De 55 a 59 years | 58 | 80 | 78 | 96 | 121 | 97 |
| De 60 a 64 years | 86 | 85 | 116 | 112 | 161 | 162 |
| De 65 a 69 years | 114 | 114 | 130 | 180 | 224 | 231 |
| De 70 a 74 years | 144 | 169 | 166 | 214 | 314 | 279 |
| De 75 a 79 years | 176 | 211 | 224 | 295 | 341 | 365 |
| De 80 years or more | 284 | 333 | 361 | 485 | 713 | 743 |
| **Total** | **988** | **1,110** | **1,248** | **1,510** | **2,052** | **1,993** |

**Table 8.** Incident cases of AF (Male) in Colombia 2010-2015 for ages. Source: SISPRO.

| **Ages** | **2010** | **2011** | **2012** | **2013** | **2014** | **2015** |
| --- | --- | --- | --- | --- | --- | --- |
| De 0 a 4 years | 1 | 2 | 3 | 1 |  |  |
| De 05 a 09 years |  |  |  |  |  | 2 |
| De 10 a 14 years | 2 | 1 |  | 1 |  | 3 |
| De 15 a 19 years | 9 | 3 | 2 | 4 | 6 | 4 |
| De 20 a 24 years | 2 | 2 | 3 | 6 | 9 | 7 |
| De 25 a 29 years | 10 | 9 | 13 | 8 | 10 | 12 |
| De 30 a 34 years | 9 | 21 | 11 | 14 | 23 | 30 |
| De 35 a 39 years | 22 | 15 | 19 | 19 | 37 | 22 |
| De 40 a 44 years | 18 | 32 | 23 | 33 | 37 | 27 |
| De 45 a 49 years | 39 | 45 | 49 | 55 | 53 | 65 |
| De 50 a 54 years | 57 | 61 | 55 | 89 | 87 | 96 |
| De 55 a 59 years | 78 | 96 | 100 | 98 | 134 | 134 |
| De 60 a 64 years | 86 | 135 | 130 | 172 | 192 | 167 |
| De 65 a 69 years | 127 | 142 | 146 | 202 | 273 | 274 |
| De 70 a 74 years | 131 | 183 | 184 | 225 | 323 | 302 |
| De 75 a 79 years | 142 | 187 | 191 | 235 | 295 | 283 |
| De 80 years or older | 163 | 217 | 252 | 309 | 485 | 484 |
| **Total** | **879** | **1,143** | **1,171** | **1,458** | **1,933** | **1,879** |

Between 2010 and 2014, 134,774,182 care services were provided by the general health social security system, for an average of 22,462,363 + 2,670,964 care services per year. The demand of care for AF in 2010 and 2015 was 0.05% and 0.10%, respectively. The demand of care for AF in women and men in 2010 was 4.2 and 3.9 per 100,000, respectively. In 2015, the demand of care for women and men was 8.1 and 7.8 per 100,000 (Table 2).

## Discussion

In our study, an AF prevalence of 0.05% was estimated, 0.38% in those older than 18 years of age, and an increase of 150% was evident between 2010 and 2015. This prevalence is lower than the global regional report, and other AF prevalence studies nationwide. The global prevalence of AF ranges from 0.4% to 2.0%, and may reach 2.5% in those older 60 years old. An increase in its prevalence is expected in the next 50 years (14,18). This study found a prevalence of 0.02% FA in patients between 40 and 50 years of age, 1.9% in patients older than 80 years of age was estimated. No differences were found by sex, except in the age group between 40-50 years old where men predominated. Globally, a prevalence of <0.5% at 40–50 years and 5–15% at 80 years has been reported, and it affects men more (14). An average of more than 2,500 new cases of AF were estimated each year, in both men and women. Globally, an incidence of 2000 per 100,000 has been reported in both sexes (18).

Regarding epidemiological studies of AF in Colombia, previous studies are scarce and have estimated higher prevalences than those reported in our study. In the study of prevalence of AF in those older than 60 years old in a population of a Colombian university hospital based on more than 2000 EKG records, a crude and adjusted prevalence of AF in those older than 60 years old was estimated at 4.8% and 3.6%, respectively. The authors of this study recognized that because the estimates came from a single source, specifically from a high complexity hospital, it does not allow the results to be generalized to the entire Colombian population (19). In the study of the prevalence of AF in a population with cardiovascular risk factors of more than 4,700 people affiliated to a healthcare provider, an AF prevalence of 0.15% was estimated. In those older than 60 years of age a prevalence of 1.6% was estimated.. This estimate is similar to that reported in the present study (20). In the cross-sectional study based on a population older than 18 years old from a specific health department, within the framework of the AFINVA registry (Actuality in Non-Valvular Atrial Fibrillation), an AF prevalence of 2.1% and 8.06% in those over 65 years of age was estimated. The authors of this study recognized that the cross-sectional design used, could influence a possible overestimation of prevalence (21).

Regarding the limitations of this study, we did not have access to the medical records and EKG records of the patients who had registered diagnosis of AF in the SISPRO database, which was a major disadvantage. This database registers only the population that has demanded services for a specific period of time, which can be a limitation when extrapolating to the entire population of the country. The adjustment based on population projections from the DANE 2005 census may influence these estimates. However, we considered that the information sources used, have improved in their completeness and quality of the data, which makes evident the information on AF in Colombia with relative strength (22).

## Conclusions

The prevalence of AF in Colombia in the population that used healthcare services is approximately 0.05% (51.4 per 100,000). Given the previous data based on cross-sectional population studies, and the limitations to generalize the results. A 66% underreporting in the diagnosis of AF can be estimated in the SISPRO database.

## Highlights

- AF is the most frequent and most important cardiac arrhythmia in the clinical practice of the adult population, both due to its incidence, and increasing age.
- AF is important for its morbidity and mortality due the ability to cause cardioembolic and cerebrovascular events.
- The global prevalence of AF is 1-2%, it increases with age and it is estimated that the risk of developing this arrhythmia throughout life after the age of 40 is 25%.
- Population studies have estimated a prevalence of AF in Colombia of 0.15%. and from 1.6% to 4.8% in those over 60 years of age.
- The prevalence of AF in Colombia of the population that used healthcare services is approximately 0.05% (51.4 per 100,000).
- An underreporting of 66% in the diagnosis of AF can be estimated in the SISPRO database.

## Data Availability

All the data produced is available online on the web pages of the Data Warehouse of the Social Protection Information System (SISPRO) upon request for access to the Colombian Ministry of Health and the National Administrative Department of Statistics (DANE) in a public way and open.

## Ethical Responsibilities

### Protection of people and animals

The authors declare that no experiments were carried out on humans or animals for this research.

### Data confidentiality

The authors declare that no patient data appears in this article.

### Right to privacy and informed consent

The authors declare that no patient data appears in this article.

